# Impact of personal protective equipment use on health care workers’ physical health during the COVID-19 pandemic: a systematic review and meta-analysis

**DOI:** 10.1101/2021.02.03.21251056

**Authors:** Petros Galanis, Irene Vraka, Despoina Fragkou, Angeliki Bilali, Daphne Kaitelidou

**Affiliations:** Faculty of Nursing, Center for Health Services Management and Evaluation, National and Kapodistrian University of Athens, Athens, Greece; Department of Radiology, P & A Kyriakou Children’s Hospital, Athens, Greece; Hospital Waste Management Unit, P & A Kyriakou Children’s Hospital, Athens, Greece

**Keywords:** adverse events, COVID-19, health care workers, personal protective equipment, physical health, SARS-CoV-2

## Abstract

**Background:** During the COVID-19 pandemic, health care workers (HCWs) caring for patients with coronavirus disease 2019 (COVID-19) in high-risk clinical settings have been obliged to wear personal protective equipment (PPE).

**Aim:** To assess the impact of PPE use on HCWs’ physical health during the COVID-19 pandemic. Also, we examined factors related with a greater risk of adverse events among HCWs due to PPE use.

**Methods:** We applied the Preferred Reporting Items for Systematic Reviews and Meta-Analysis guidelines and the Cochrane criteria for this systematic review and meta-analysis. We searched PubMed, Medline, Scopus, ProQuest, CINAHL and pre-print services (medRxiv) from January 1, 2020 to December 27, 2020.

**Findings:** Our review included 14 studies with 11,746 HCWs from 16 countries. The estimated overall prevalence of adverse events among HCWs was 78% (95% CI: 66.7-87.5%) with a range from 42.8% to 95.1% among studies. The prevalence of adverse events was higher for the studies with poor quality compared to those with moderate quality (83.5% vs. 67.1%), while increased sample size was related with decreased prevalence (p<0.001). The most frequent adverse events were headache (55.9%), dry skin (54.4%), dyspnoea (53.4%), pressure injuries (40.4%), itching (39.8%), hyperhidrosis (38.5%), and dermatitis (31.0%). Among others, the following factors were related with the risk of adverse events among HCWs due to PPE use: female gender, younger age, obesity, diabetes mellitus, smoking pre-existing headache, longer duration of shifts wearing PPE, increased consecutive days with PPE, and increased exposure to confirmed or suspected COVID-19 patients.

**Conclusion:** The frequency of adverse events amongst HCWs due to PPE use is very high. Further studies should be conducted since the limitations of this review do not allow us to infer conclusive results especially in case of risk factors for the occurrence of adverse events. Healthcare facilities should take the necessary precautions and change the working conditions during the COVID-19 pandemic to prevent adverse events associated with PPE use and minimize harm to HCWs.

## Introduction

Health care workers (HCWs) can be exposed to the severe acute respiratory syndrome coronavirus 2 (SARS-CoV-2) through clinical settings or community transmission and are essential workers at risk for coronavirus disease 2019 (COVID-19). According to Centers for Disease Control and Prevention (CDC) during February 12-July 16, 2020, in the USA, 11% of patients had been identified as HCWs [1], while during March 1-May 31, 2020, among hospitalized adults, 5.9% were HCWs [2]. A meta-analysis [3] found that the prevalence of hospitalization among HCWs infected with COVID-19 is 15.1% and the mortality is 1.5%, while another meta-analysis [4] found that the proportion of SARS-CoV-2 positive HCWs among all COVID-19 patients is 10.1% and the mortality is significantly lower in HCWs as compared to that of all patients (0.3% vs. 2.3%). According to an analysis included studies only in Australia between January 25^th^ and July 8^th^, HCWs were 2.69 times more likely to contract COVID-19 than the general population [5]. Also, the seroprevalence of SARS-CoV-2 antibodies among HCWs is high (8.7%) especially in North America (12.7%) compared to Europe (8.5%), Africa (8.2), and Asia (4%) [6].

During the COVID-19 pandemic, HCWs caring for patients with COVID-19 in high-risk clinical settings such as isolation wards, intensive care units, emergency rooms and general medical wards have been obliged to wear personal protective equipment (PPE). PPE includes equipment or specific clothing (e.g. goggles, mop caps, respirator masks, face shields, shoe covers, gowns and gloves) that protects HCWs against infectious materials [7]. The necessity of PPE to prevent transmission of viruses to HCWs has already proven during the severe acute respiratory distress syndrome (SARS) [8] and Ebola epidemic [9]. During the COVID-19 pandemic, HCWs have to wear PPE unceasingly for more than 6-8 hours in a shift. Moreover, inappropriate PPE reuse (e.g. donning of a used PPE item without contamination) due to global PPE shortages remains affecting HCWs and patients safety and the sustainability of health care systems [10–13]. Under these circumstances, World Health Organization diffuses recommendations for optimizing PPE use by HCWs caring for suspected or confirmed COVID-19 patients especially in countries with severe PPE shortages [7].

Several studies have already shown that adverse reactions from PPE use amongst HCWs are common including dermatitis, allergy, atopy, facial itch, acne, rash etc. [14–18]. Considering the long-time wearing of PPE among HCWs and PPE shortages during the COVID-19 pandemic, we anticipated a high incidence of physical health problems due to PPE use among HCWs. To our knowledge, the overall impact of PPE use on HCWs’ physical health during COVID-19 pandemic is unknown. Thus, the primary aim of this systematic review and meta-analysis was to assess the impact of PPE use on HCWs’ physical health during COVID-19 pandemic. The secondary objective was to examine factors related with a greater risk of adverse events among HCWs due to PPE use.

## Methods

### Data sources and strategy

We applied the Preferred Reporting Items for Systematic Reviews and Meta-Analysis (PRISMA) guidelines [19] and the Cochrane criteria [20] for this systematic review and meta-analysis. We searched PubMed, Medline, Scopus, ProQuest, CINAHL and pre-print services (medRxiv) from January 1, 2020 to December 27, 2020. Also, we examined reference lists of all relevant articles and we removed duplicates. We applied the following filters during the search in the databases: humans, English language, and journal article. We used the following strategy searching in title/abstract query: ((“health care worker*” OR “healthcare worker*” OR “healthcare personnel” OR “health care personnel” OR “health personnel” OR “health care professional*” OR “healthcare professional*” OR staff OR “nursing staff” OR professional* OR worker* OR doctor* OR physician* OR clinician* OR nurs* OR midwives OR midwife* OR paramedic* OR practitioner*) AND (“personal protective equipment”)) AND (COVID-19 OR COVID19 OR COVID OR SARS-CoV* OR “Severe Acute Respiratory Syndrome Coronavirus*” OR coronavirus*). The study protocol was registered with PROSPERO (CRD42021228221).

### Selection and eligibility criteria

Two independent reviewers performed study selection and discrepancies were resolved by a third, senior reviewer. We initially screened title and abstract of the records and then full-text. We included studies that examine impact of PPE use on HCWs’ physical health during COVID-19 pandemic. Also, we included studies examining factors related with a greater risk of adverse events among HCWs due to PPE use. We examined articles that were published in English, except reviews, qualitative studies, protocols, case reports, editorials, and letters to the Editor. All types of HCWs directly involving in the management of COVID-19 patients were accepted for inclusion, while we excluded studies with health care students and general population. Also, we excluded studies that examined effects of PPE use on psychological or mental health of HCWs.

### Data extraction and quality assessment

We extracted the following data from each study: authors, location, sample size, age, gender, study design, sampling method, assessment of the adverse events, response rate, data collection time, type of publication (journal or pre-print service), number and type of adverse events among HCWs, factors related with a greater risk of adverse events, and the level of analysis (univariate or multivariable).

Two reviewers used the Joanna Briggs Institute critical appraisal tools to assess quality of studies as poor, moderate or good [21]. Regarding cross-sectional studies, an 8-point scale is used with a score of ≤3 refers to poor quality, a score of 4-6 points refers to moderate quality, and a score of 7-8 points refers to good quality.

### Statistical analysis

For each study we extracted the sample size and adverse events that occurred among HCWs due to PPE use. We initially calculated the prevalence of any adverse event and the 95% confidence interval (CI) for each included study. Then, we transformed these prevalences with the Freeman-Tukey Double Arcsine method before pooling [22]. Moreover, we pooled the results for adverse events that occurred among HCWs at least in three studies. We assessed between-studies heterogeneity with the Hedges Q statistic and I^2^ statistics. I^2^ values higher than 75% indicates high heterogeneity, while a p-value<0.1 for the Hedges Q statistic indicates statistically significant heterogeneity [23]. A random effect model was applied to estimate pooled effects since the heterogeneity between results was very high [23]. A leave-one-out sensitivity analysis was performed to determine the influence of each study on the overall effect. We used a funnel plot and the Egger’s test to assess the publication bias with a P-value<0.05 indicating publication bias [24]. A priori, we considered gender, age, sample size, the continent that studies were conducted, studies quality, study design, assessment of the outcome, data collection time, and publication type (journal or pre-print service) as sources of heterogeneity. Due to the limited data and limited variability of some of these variables, we decided to perform meta-regression analysis and subgroup analysis considering gender, sample size, studies quality, and data collection time as sources of heterogeneity. We did not perform meta-analysis for the factors related with the occurrence of adverse events among HCWs since the data were very limited and highly heterogeneous. We used the OpenMeta[Analyst] to perform meta-analysis [25].

## Results

### Identification and selection of studies

Flowchart of the literature search is summarized in PRISMA format and it is shown in Figure 1. We initially identified 2699 potential records through PubMed, Medline, Scopus, ProQuest, CINAHL and medRxiv removing duplicates. After the screening of the titles and abstracts, we removed 2671 records and we added 4 more records found by the reference lists scanning. Finally, we included 14 studies [26–39] in this meta-analysis that met our inclusion criteria.

**Figure 1.**
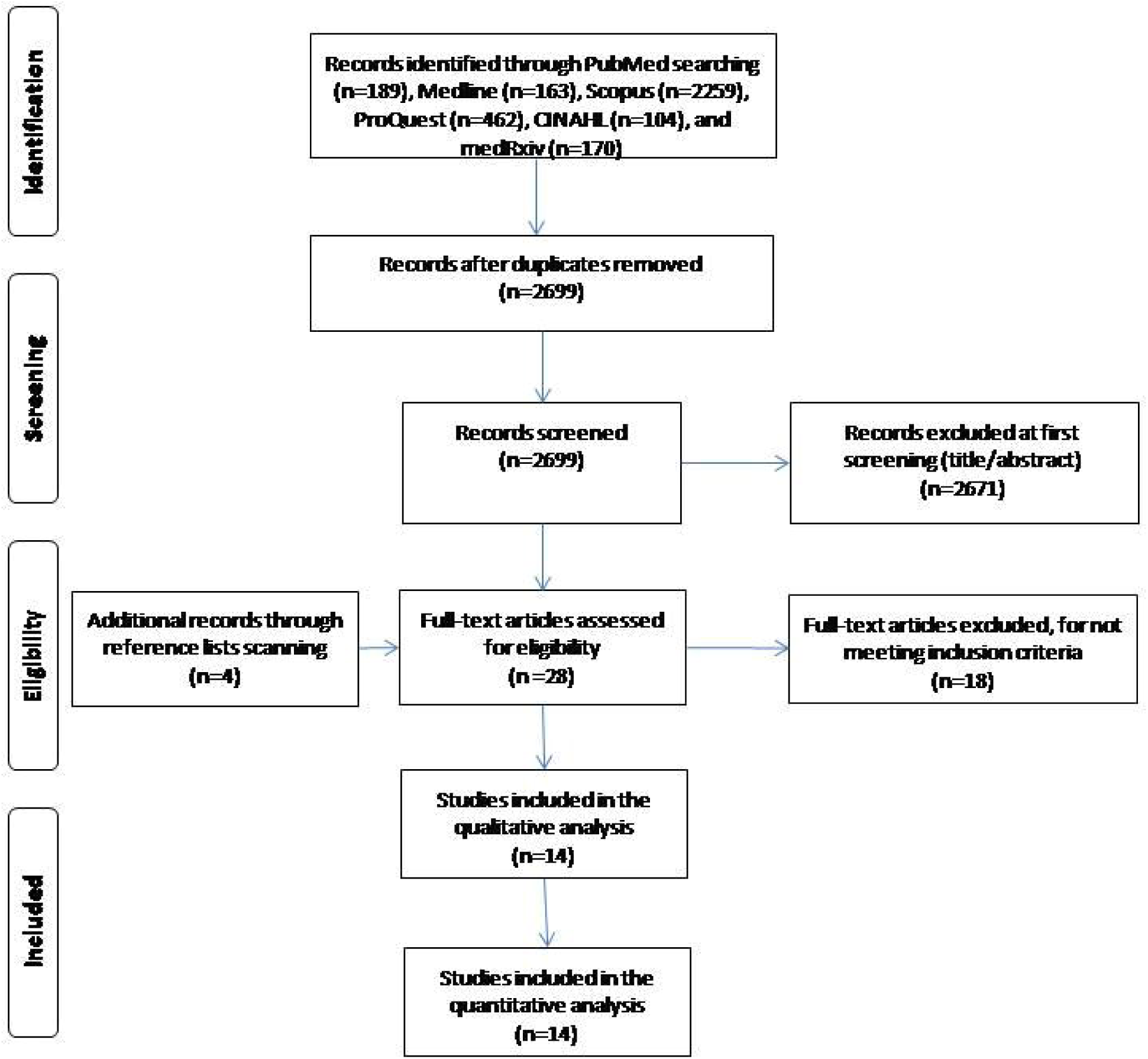
Flowchart of the literature search according to the Preferred Reporting Items for Systematic Reviews and Meta-Analysis.

### Characteristics of the studies

Main characteristics of the studies included in our systematic review and meta-analysis are shown in Table 1. A total of 11,746 HCWs from 16 countries were included in this review. Number of HCWs in studies ranged from 40 to 4306, while females’ percentage ranged from 46.0% to 91.8%. The majority of studies were conducted in Asia (n=10) [26,28,30–33,35,36,38,39], two studies were conducted in Europe [34,37], one study was conducted in South America [27], and one study included HCWs from 10 countries [29]. All studies were cross-sectional, while 13 studies [26–30,32–39] used a convenience sample method and one study [31] used a purposeful sampling method. Assessment of adverse events was self-reported through questionnaires in 13 studies [26–35,37–39], while in one study [36] a clinical diagnosis was performed. All studies were published in journals and seven studies [26,28,29,31,32,38,39] reported response rate.

**Table 1.**
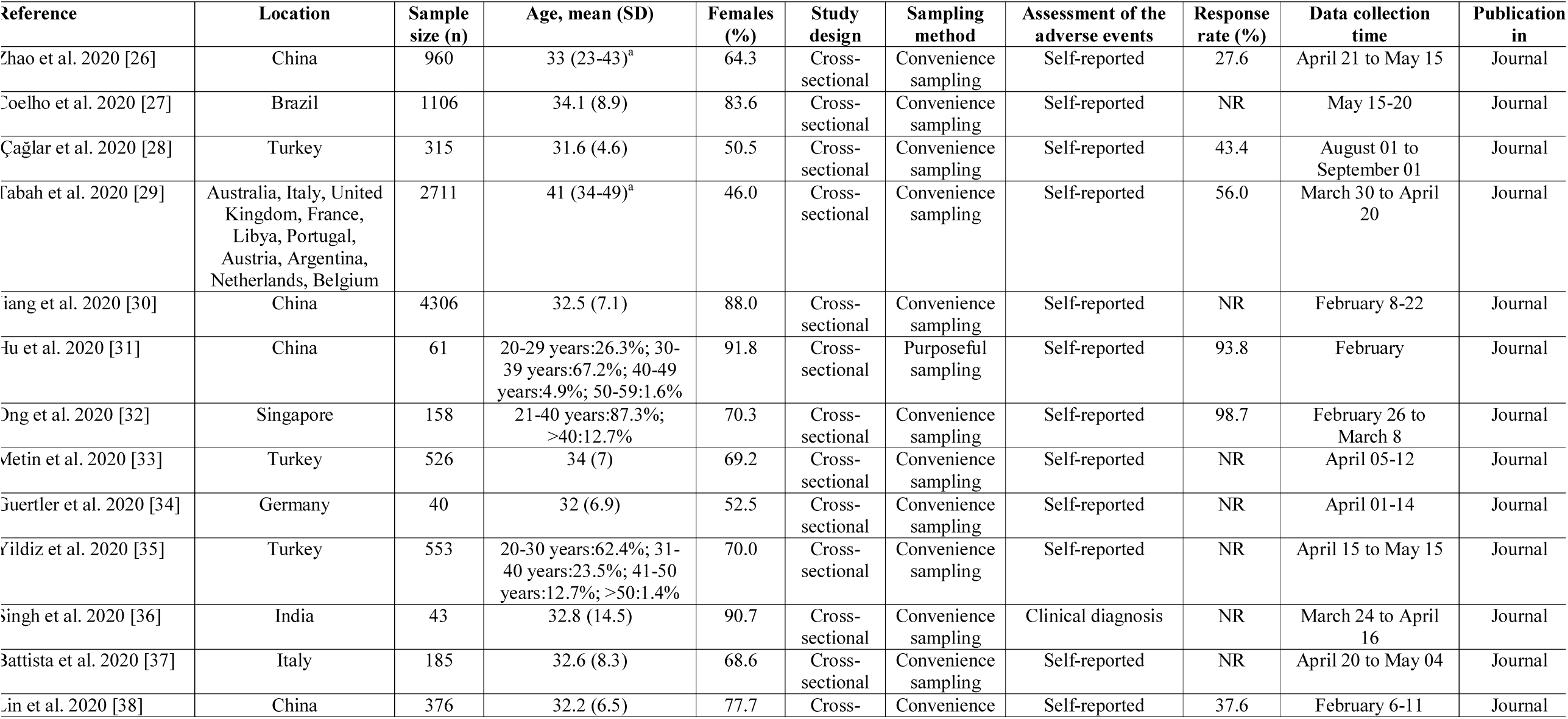
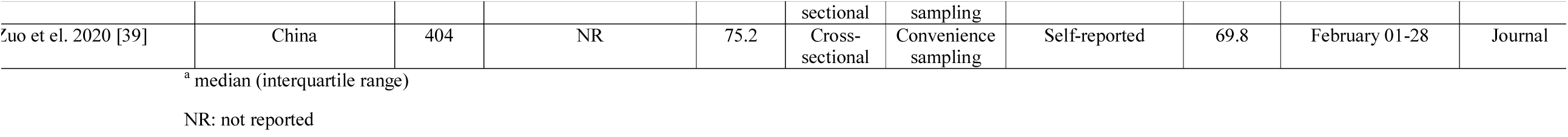
Main characteristics of the studies included in this systematic review.

### Quality assessment

Quality assessment of cross-sectional studies included in this systematic review is shown in Table 2. Quality was poor in nine studies [26,29,31,33–37,39] and moderate in five studies [27,28,30,32,38].

**Table 2a.** Quality of cross-sectional studies included in this systematic review.

|  | Zhao<br>et al.<br>2020<br>[26] | Coelho<br>et al.<br>2020<br>[27] | Çağlar et<br>al. 2020<br>[28] | Tabah et<br>al. 2020<br>[29] | Jiang et<br>al. 2020<br>[30] | Hu et<br>al.<br>2020<br>[31] | Ong et<br>al. 2020<br>[32] | Metin et<br>al. 2020<br>[33] | Guertler<br>et al.<br>2020<br>[34] | Yildiz et<br>al. 2020<br>[35] |
| --- | --- | --- | --- | --- | --- | --- | --- | --- | --- | --- |
| 1. Were the criteria for inclusion in the sample clearly defined? | X | X | X |  | X |  | X |  |  |  |
| 2. Were the study subjects and the setting described in detail? | X | X | X | X | X |  | X | X | X | X |
| 3. Was the exposure measured in a valid and reliable way? |  |  |  |  |  |  |  |  |  |  |
| 4. Were objective, standard criteria used for measurement of the condition? |  |  |  |  |  |  |  |  |  |  |
| 5. Were confounding factors identified? |  | X | X |  | X |  | X |  |  |  |
| 6. Were strategies to deal with confounding factors stated? |  | X | X |  | X |  | X |  |  |  |
| 7. Were the outcomes measured in a valid and reliable way? |  |  |  |  |  |  |  |  |  |  |
| 8. Was appropriate statistical analysis used? | X | X | X | X | X | X | X | X | X | X |
| **Total quality** | Poor | Moderate | Moderate | Poor | Moderate | Poor | Moderate | Poor | Poor | Poor |

**Table 2b.**
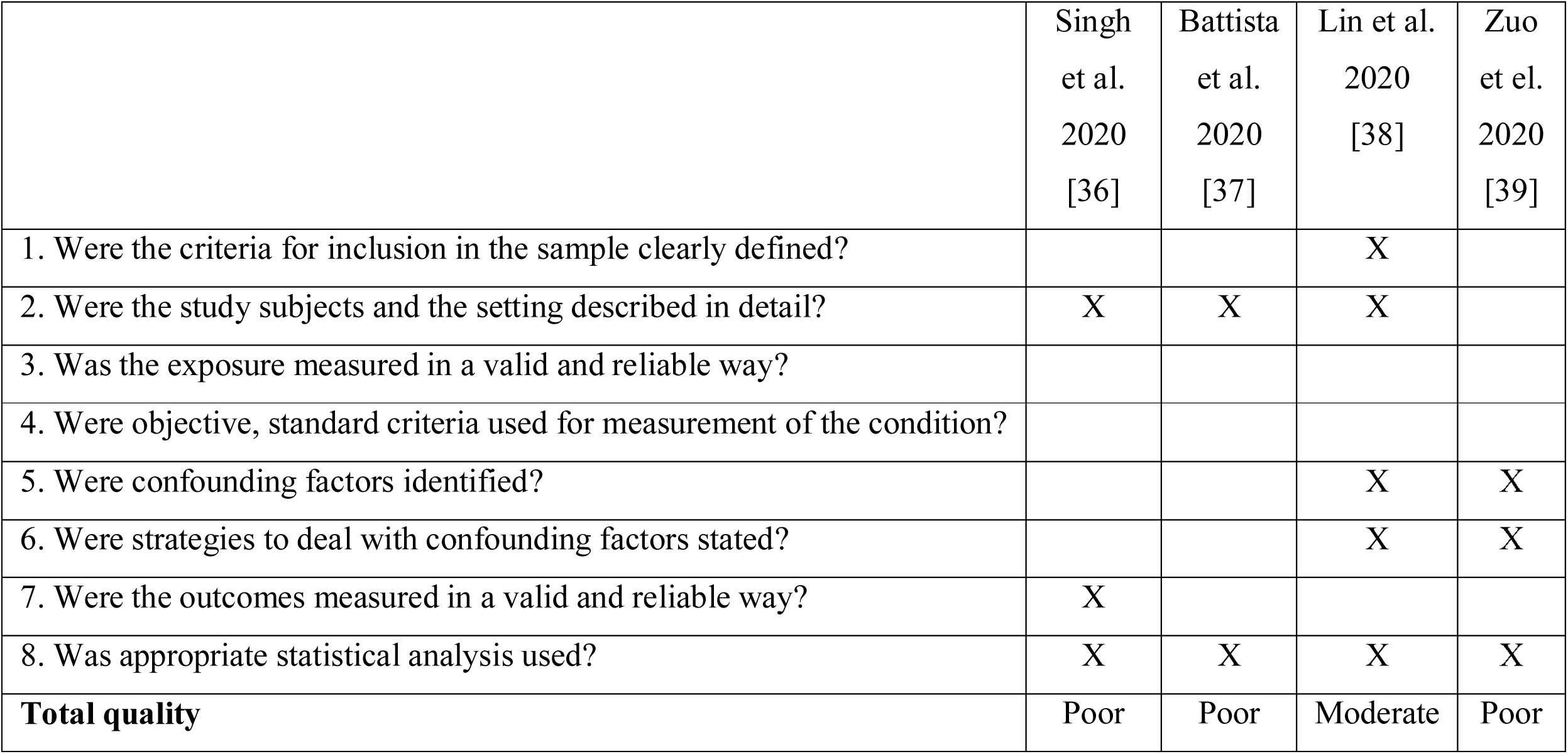
Quality of cross-sectional studies included in this systematic review.

### Meta-analysis

A random effect model was applied to estimate pooled prevalence of adverse events since the heterogeneity between results was very high (I^2^=99.39, p-value for the Hedges Q statistic < 0.001). The estimated overall prevalence of adverse events among HCWs was 78% (95% CI: 66.7-87.5%) (Figure 2). Prevalence among studies ranged from 42.8% [30] to 95.1% [31]. A leave-one-out sensitivity analysis showed that no single study had a disproportional effect on the pooled prevalence, which varied between 76.4% (95% CI: 64.5-86.4%), with Hu et al. [31] excluded, and 80.3% (95% CI: 73.8-86.1%) with Jiang et al. [30] excluded. A publication bias was potential since p-value for Egger’s test was <0.05 and the shape of the funnel plot was asymmetrical (Web Figure 2).

**Figure 2.**
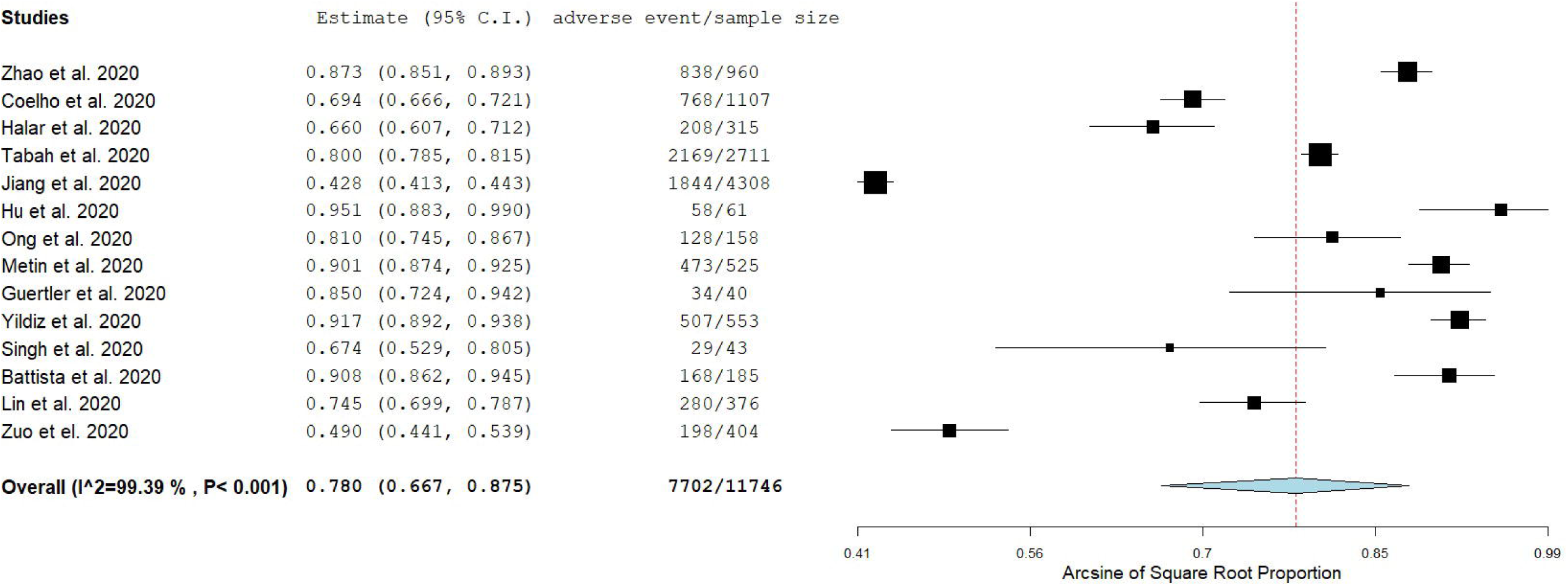
Forest plot of the prevalence of adverse events among health care workers.

According to subgroup analysis, the prevalence of adverse events was higher for the studies with poor quality (83.5% [95% CI: 75.4-90.2%], I^2^=97.64) compared to those with moderate quality (67.1% [95% CI: 50.4-81.8%], I^2^=99.13). Meta-regression analysis identified that increased sample size was related with decreased prevalence of adverse events among HCWs (p<0.001), (Web figure 3). Also, the prevalence of adverse events was independent of the gender distribution (p=0.32), and data collection time (p=0.63).

Adverse events among HCWs due to personal protective equipment use during COVID-19 pandemic are listed in Table 3. We pooled the results for adverse events that occurred among HCWs at least in three studies and the results are presented in Table 4. According to the pooled results, the adverse events that occurred more often were headache (55.9% [95% CI: 35.8-75.0%]), dry skin (54.4% [95% CI: 25.4-81.8%]), dyspnoea (53.4% [95% CI: 27.2-78.6%]), pressure injuries (40.4% [95% CI: 27.7-53.8%]), itching (39.8% [95% CI: 16.2-66.3%]), hyperhidrosis (38.5% [95% CI: 15.3-64.9%]), and dermatitis (31.0% [95% CI: 11.1-55.5%]).

**Table 3a.** Adverse events among health care workers due to personal protective equipment use during the COVID-19 pandemic in the studies included in this systematic review.

| **Reference** | **Any adverse event** | **Dry skin** | **Pressure injuries** | **Headache** | **Dermatitis** | **Allergy** | **Rash** | **Itching** | **Pain** |
| --- | --- | --- | --- | --- | --- | --- | --- | --- | --- |
| Zhao et al. 2020 [26] | 838 (87.3) | 199 (20.7) |  | 516 (53.8) | 146 (15.2) | 162 (16.9) | 222 (23.1) |  |  |
| Coelho et al. 2020 [27] | 768 (69.4) |  | 768 (69.4) |  |  |  |  |  |  |
| Çağlar et al. 2020 [28] | 208 (66.0) |  |  | 115 (36.5) | 64 (20.3) |  |  |  |  |
| Tabah et al. 2020 [29] | 2169 (80.0) |  | 1088 (40.1) | 696 (25.7) |  |  |  |  |  |
| Jiang et al. 2020 [30] | 1844 (42.8) |  | 1293 (30.0) |  |  |  |  |  |  |
| Hu et al. 2020 [31] | 58 (95.1) | 15 (24.6) | 42 (68.9) |  |  | 7 (11.5) | 10 (16.4) | 17 (27.9) | 7 (11.5) |
| Ong et al. 2020 [32] | 128 (81.0) |  |  | 128 (81.0) |  |  |  |  |  |
| Metin et al. 2020 [33] | 473 (90.1) | 473 (90.1) |  |  | 379 (72.5) | 100 (19.1) | 100 (19.1) |  |  |
| Guertler et al. 2020 [34] | 34 (85.0) | 32 (82.1) | 13 (32.5) |  | 6 (15.0) | 8 (20.5) |  | 12 (30) |  |
| Yildiz et al. 2020 [35] | 507 (91.7) | 507 (91.7) |  | 410 (74.1) |  |  |  |  | 494 (89.3) |
| Singh et al. 2020 [36] | 29 (67.4) | 16 (37.2) | 11 (25.6) | 27 (62.8) | 17 (39.5) | 3 (7.0) | 21 (48.8) | 29 (67.4) |  |
| Battista et al. 2020 [37] | 168 (90.8) |  |  |  | 54 (29.2) |  |  | 118 (63.8) | 93 (50.3) |
| Lin et al. 2020 [38] | 280 (74.5) | 258 (68.6) |  |  |  |  |  |  |  |
| Zuo et al. 2020 [39] | 198 (49.0) | 47 (11.6) | 76 (18.8) |  |  |  | 50 (12.4) | 60 (14.9) | 13 (3.2) |

Values are expressed as n (%)

**Table 3b.**
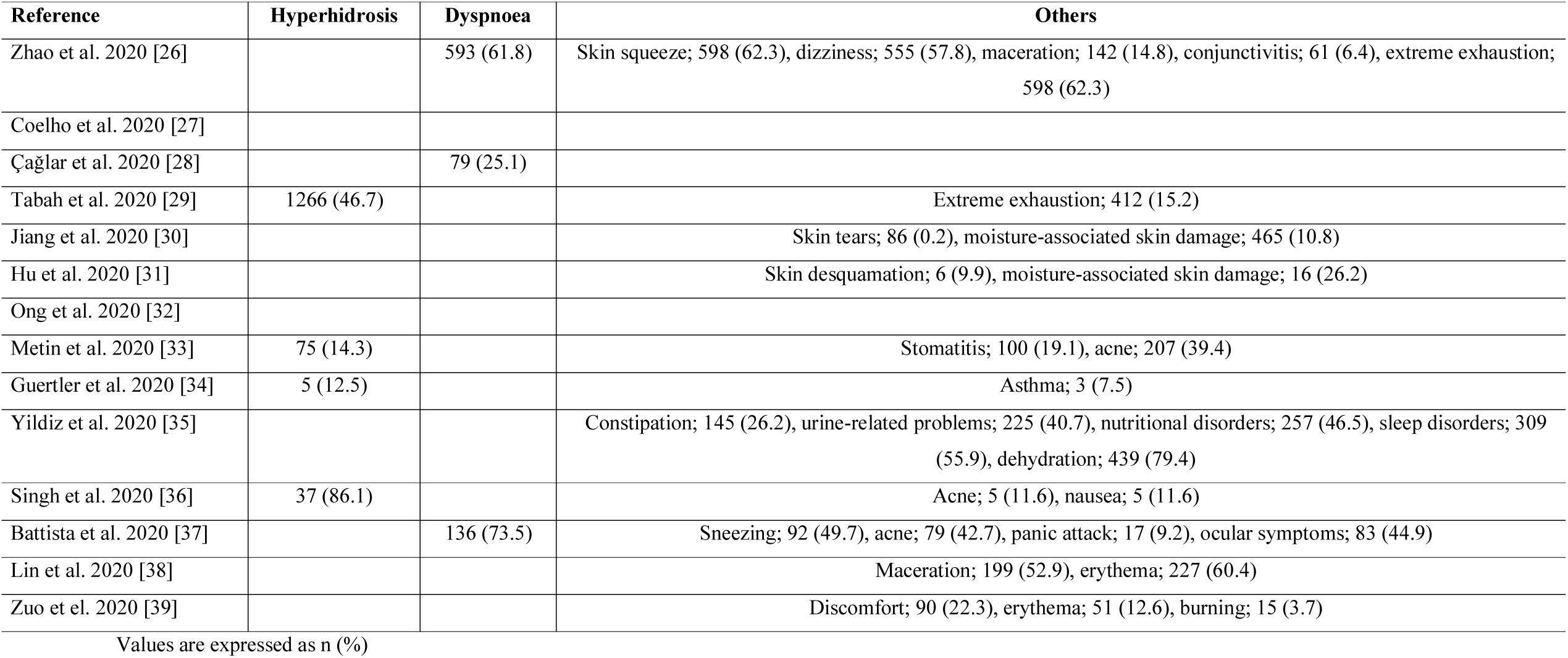
Adverse events among health care workers due to personal protective equipment use during the COVID-19 pandemic in the studies included in this systematic review.

| Reference | Hyperhidrosis | Dyspnoea | Others |
| --- | --- | --- | --- |
| Zhao et al. 2020 [26] |  | 593 (61.8) | Skin squeeze; 598 (62.3), dizziness; 555 (57.8), maceration; 142 (14.8), conjunctivitis; 61 (6.4), extreme exhaustion; 598 (62.3) |
| Coelho et al. 2020 [27] |  |  |  |
| Çağlar et al. 2020 [28] |  | 79 (25.1) |  |
| Tabah et al. 2020 [29] | 1266 (46.7) |  | Extreme exhaustion; 412 (15.2) |
| Jiang et al. 2020 [30] |  |  | Skin tears; 86 (0.2), moisture-associated skin damage; 465 (10.8) |
| Hu et al. 2020 [31] |  |  | Skin desquamation; 6 (9.9), moisture-associated skin damage; 16 (26.2) |
| Ong et al. 2020 [32] |  |  |  |
| Metin et al. 2020 [33] | 75 (14.3) |  | Stomatitis; 100 (19.1), acne; 207 (39.4) |
| Guertler et al. 2020 [34] | 5 (12.5) |  | Asthma; 3 (7.5) |
| Yildiz et al. 2020 [35] |  |  | Constipation; 145 (26.2), urine-related problems; 225 (40.7), nutritional disorders; 257 (46.5), sleep disorders; 309 (55.9), dehydration; 439 (79.4) |
| Singh et al. 2020 [36] | 37 (86.1) |  | Acne; 5 (11.6), nausea; 5 (11.6) |
| Battista et al. 2020 [37] |  | 136 (73.5) | Sneezing; 92 (49.7), acne; 79 (42.7), panic attack; 17 (9.2), ocular symptoms; 83 (44.9) |
| Lin et al. 2020 [38] |  |  | Maceration; 199 (52.9), erythema; 227 (60.4) |
| Zuo et el. 2020 [39] |  |  | Discomfort; 90 (22.3), erythema; 51 (12.6), burning; 15 (3.7) |

Values are expressed as n (%)

**Table 4.** Meta-analysis for the adverse events among health care workers due to personal protective equipment use during the COVID-19 pandemic.

| **Adverse event** | **No. of studies** | **Pooled prevalence** | **95% confidence interval** | **I<sup>2</sup>** | **P-value for the Hedges Q statistic** |
| --- | --- | --- | --- | --- | --- |
| Dry skin | 8 | 54.4 | 25.4 – 81.8 | 99.60 | <0.001 |
| Pressure injuries | 7 | 40.4 | 27.7 – 53.8 | 99.12 | <0.001 |
| Headache | 6 | 55.9 | 35.8 – 75.0 | 99.32 | <0.001 |
| Dermatitis | 6 | 31.0 | 11.1 – 55.5 | 99.09 | <0.001 |
| Allergy | 5 | 16.4 | 13.2 – 19.8 | 47.76 | 0.105 |
| Rash | 5 | 21.4 | 15.1 – 28.5 | 90.04 | <0.001 |
| Itching | 5 | 39.8 | 16.2 – 66.3 | 97.62 | <0.001 |
| Pain | 4 | 35.5 | 0.3 – 88.1 | 99.73 | <0.001 |
| Hyperhidrosis | 4 | 38.5 | 15.3 – 64.9 | 98.98 | <0.001 |
| Dyspnoea | 3 | 53.4 | 27.2 – 78.6 | 98.81 | <0.001 |

### Risk factors for adverse events

Eleven studies [26–33,35,38,39] investigated risk factors for adverse events among HCWs due to personal protective equipment use during COVID-19 pandemic (Table 5). Six studies [21,27,28,32,38,39] used multivariable models to eliminate confounding factors, while all studies except one [33] measured the occurrence of any adverse event as the dependent variable.

**Table 5.**
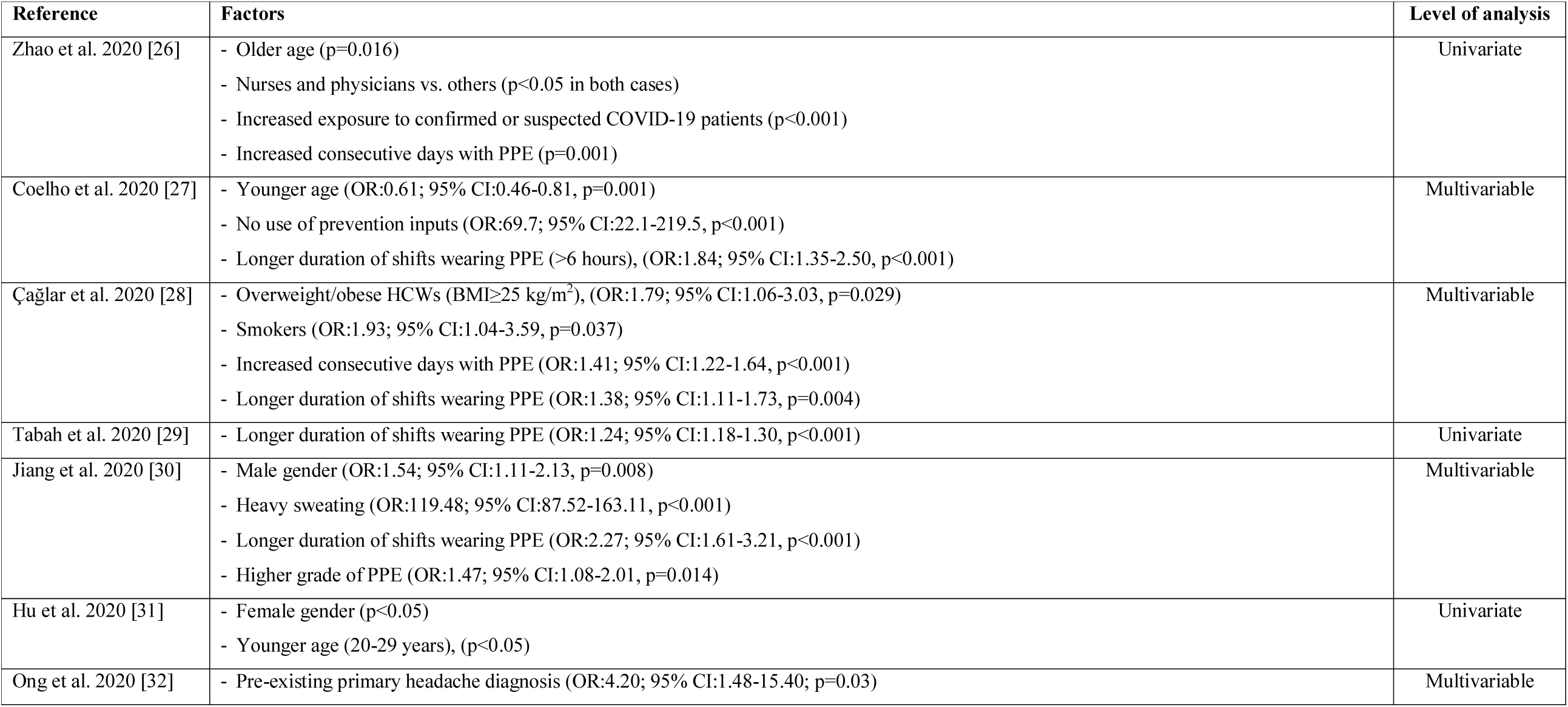
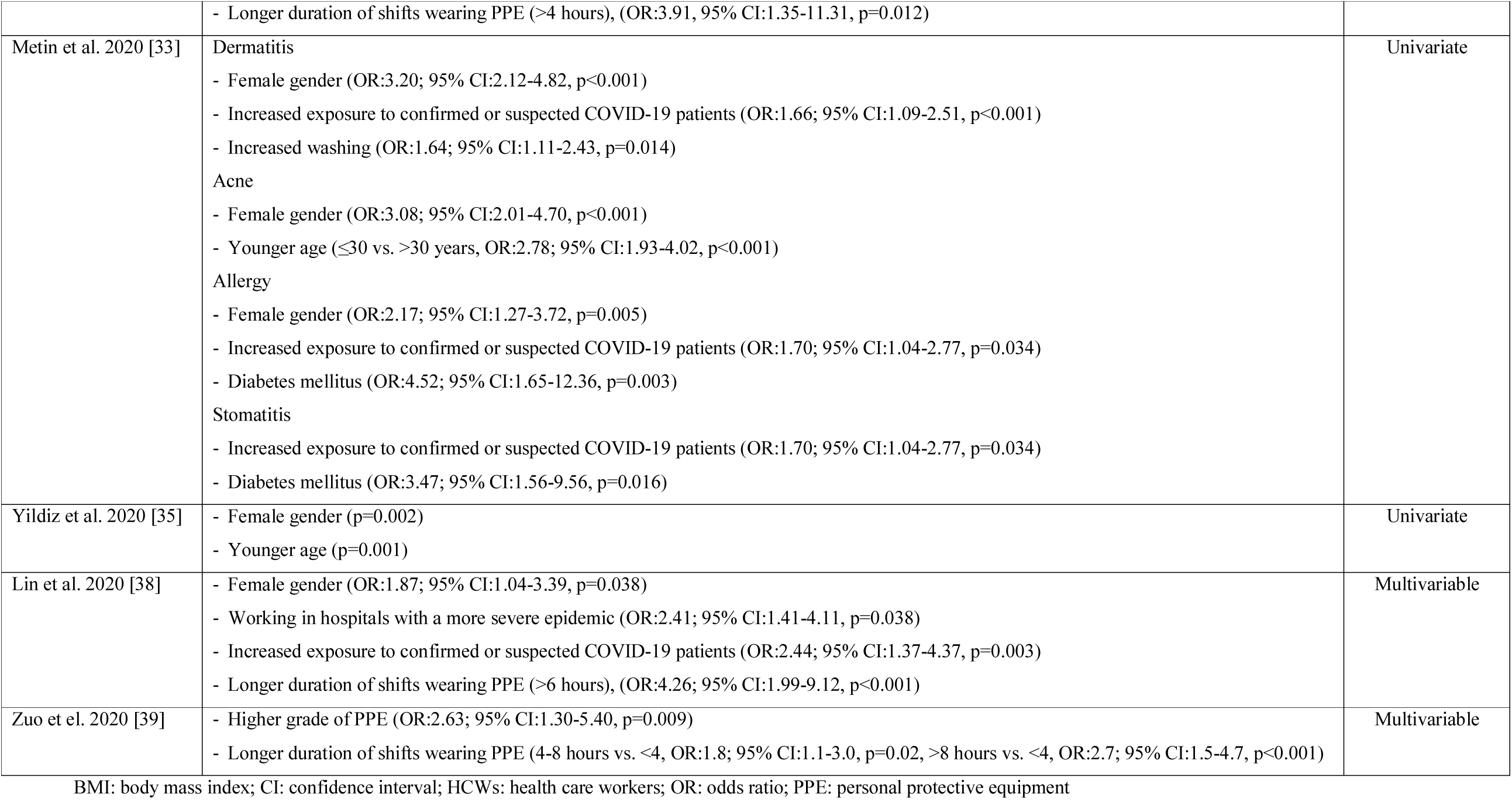
Factors related with a greater risk of adverse events among health care workers due to personal protective equipment use during the COVID-19 pandemic in the studies included in this systematic review.

We found that demographic, clinical and job characteristics were related with the risk of adverse events among HCWs due to PPE use.

Regarding gender, four studies [31,33,35,38] found that females had higher risk of adverse events with ORs ranging from 1.87 to 3.20, while one study [30] found the opposite (OR:1.54 for males). Moreover, four studies [27,31,33,35] showed that younger age was associated with increased risk of adverse events, while one study [26] showed the opposite. Among HCWs, nurses and physicians were at the greater risk of developing adverse events [26].

Several clinical characteristics of the HCWs affected the occurrence of adverse events. In particular, comorbidity such as diabetes mellitus, obesity, pre-existing headache and smoking significantly increased risk of adverse events [28,32,33]. Similar, heavy sweating was a risk factor for adverse events [30].

We found that job characteristics affected adverse events in a significant way. The longer duration of shifts wearing PPE, the greater the risk of adverse events with ORs ranging from 1.24 to 4.26 [27–30,32,38]. Two studies [27,38] found that shifts >6 hours was a risk factor, while two studies [32,39] found a different cut-off point of 4 hours. Moreover, increased consecutive days with PPE [26,28] and higher grade of PPE [30,39] significantly increased risk of adverse events among HCWs. Our review showed that increased exposure to confirmed or suspected COVID-19 patients [26,33,38], working in hospitals with a more severe epidemic [38], and no use of prevention inputs [27] increased the probability of adverse events.

## Discussion

To our knowledge, this is the first systematic review and meta-analysis that investigates the impact of PPE use on HCWs’ physical health during the COVID-19 pandemic. Also, we searched for risk factors related with adverse events among HCWs.

We found that the overall prevalence of adverse events among HCWs was very high (78%) with a wide range from 42.8% to 95.1% among studies. PPE use amongst HCWs is related with skin reactions such as dermatitis, allergy, atopy, facial itch, acne, rash [14–18]. HCWs wear PPE items for long periods of time due to the shortage of PPE especially at the beginning of the COVID-19 pandemic and the increased workload in healthcare facilities [40,41]. This scenario increases considerably the risk of adverse events such as skin reactions. The problem is further complicated by the lack of training and awareness among HCWs about the use of PPE [42,43]. During the COVID-19 pandemic, HCWs have to encounter several challenges regarding PPE such as donning (putting on) and doffing (taking off) equipment in the appropriate way, wearing PPE items for long periods of time, difficulties in communication with patients and colleagues etc. [44,45]. HCWs should undergo compulsory training on the correct use of PPE and guidelines should emphasize on the correct use of PPE using video training and simulations than traditional methods of teaching [7,45].

According to our results, the most prevalent physical complaint from the use of PPE was headaches. Previous studies confirm that headaches are common among HCWs when the N95 face mask is used especially for a prolonged period [46–48]. It is well known that headaches could arise from the continuous pressure of pericranial soft issues by putting on objects with tight straps around the head, e.g. helmets, hats, goggles [49–51]. Also, breathing discomfort due to N95 face mask has also been reported in the literature confirming our finding that dyspnoea is a common adverse event among HCWs due to PPE use [52–54]. A survey among dental professionals during the COVID-19 pandemic found that the prolonged use of filtering facepiece 2 (FFP2) respirators was related with moderate breathing difficulties [55]. Moreover, increased levels of anxiety and stress among HCWs during the pandemic [56,57] may contribute to breathing difficulties.

We found that skin reactions (e.g. dry skin, itching, dermatitis and rash) were the more frequent adverse events that HCWs encountered. While increased use of gloves and masks and excessive sanitizing of hands amongst HCWs are indispensable to prevent transmission of SARS-CoV-2, they also has negative implications leading to a removal of normal bacterial flora and a disruption of the natural protective skin barrier [58–60]. In that case, the frequency and the severity of occupational skin diseases increase [61–63].

Adverse events caused by PPE use are a comprehensive effect with sociodemographic, clinical and job characteristics as the contributing factors. Regarding the sociodemographic factors, we found that gender, age and type of occupation affect the impact of PPE use on HCWs’ physical health. The effect of gender and age is controversial. In particular, four studies [31,33,35,38] found that adverse events are more common among females and one study [30] found the opposite. A multi-center survey in China [64] found a higher prevalence of pressure injuries in male hospitalized patients while another study with outpatients in Turkey [65] found that acne, hand eczema and urticaria are more common in females and seborrheic dermatitis is more common in males. Differences in hormones, genetic factors, activity levels, hygiene behavior and use of skin care products could explain differences in skin reactions among males and females HCWs. Regarding age, four studies [27,31,33,35] found that younger age is related with greater risk of skin reactions, while one study [26] found the opposite. Several studies found that skin reactions are more frequent in young adults [65–67].

According to our review, comorbidity is a risk factor on new-onset symptoms from the PPE use. In particular, obesity, smoking, diabetes mellitus, and pre-existing headache were related with increased odds of adverse events. Obesity and smoking decrease cardiopulmonary capacity causing dyspnea [68,69]. Obese individuals and smokers could face more symptoms because of the use of masks without valve that brings difficulties in breathing. Laferty and McKay [70] found that N95 masks cause breathing resistance resulting on a decrease in SpO_2_ and an increase in CO_2_ levels. Moreover, isolation gowns cover the entire body causing heavy sweating and continuous dehydration especially among smokers and obese individuals. A scoping review [55] among dental professionals has revealed moderate breathing difficulties due to the use of filtering facepiece 2 respirators, while the prolonged duration of respirators usage was related with headaches. This finding is confirmed by a study [46] that was conducted during the SARS pandemic and found that 37.3% of HCWs who were N95 face-masks developed headaches. This percentage was even higher (81%) in a study [32] that was conducted during the COVID-19 pandemic and found also that the odds of headache were 4.2 times higher in HCWs with pre-existing headache than among those without a pre-existing headache. Likelihood of developing headache was greater among HCWs with a long-term utilization of N95 face masks [32]. Prolonged use of N95 masks could result in hypercapnia and hypoxemia which led to headache [31].

Seven studies [27–30,32,38,39] in our review found that the duration of PPE use is an important risk factor for adverse events among HCWs. The literature comes to an agreement with this finding since Lim et al. [46] during the SARS pandemic revealed that the increased duration of N95 mask use is related with headaches development, while Shenal et al. [48] found a relation between prolonged wear of respiratory protection and discomfort. Also, longer wearing time of N95 respirators, surgical masks and goggles compress cheeks, ears, nose bridge, and forehead which could be the main cause of skin and pressure injuries on the head and face [71]. Additionally, the longer the wearing time of PPE items, the more the sweaty with heavy sweating stimulates the skin causing redness, itching and pain [30]. The problem is further complicated by the increased consecutive days with PPE leading to more adverse skin effects among HCWs [26,28]. Moreover, increased exposure to confirmed or suspected COVID-19 patients is related with increased wearing time of PPE because of the contagiousness of SARS-CoV-2 [26,33,38]. In light of the above situations, daily wearing time of PPE items among HCWs should be decreased to protect them and avoid the adverse impact of PPE use.

Our study has several limitations introducing bias. First, 10 out of 14 included studies were conducted in Asia and thus further studies should be performed worldwide, allowing us to generalize the results. Also, quality of studies was poor (in nine studies) or moderate (in five studies), while adverse events were more frequent in studies with poor quality compared to those with moderate quality. There is a need to perform more valid studies since studies with poor quality may inflate the results. In the same way, the fact that the assessment of adverse events was self-reported in 13 out of 14 studies may introduce information bias that exaggerates the frequency of adverse events. This bias could be eliminated with clinical diagnosis of adverse events due to PPE use. Variability in study designs and populations introduces high heterogeneity in our meta-analysis. We applied a random effect model and we performed subgroup and meta-regression analysis to overcome this issue. We searched six databases and the reference lists of the studies included in our review but always there is a probability to omit relevant studies. Data regarding the factors that were related with a greater risk of adverse events were scarce and only six studies used multivariable analysis to eliminate confounders. Also, causal inferences between risk factors and adverse events are impossible since all studies were cross-sectional. Thus, studies with more appropriate design (e.g. cohort studies and case-control studies) and more sophisticated analysis should be conducted to infer more valid results regarding risk factors for adverse events due to PPE use.

In conclusion, the frequency of adverse events among HCWs due to PPE use is very high, while there are several sociodemographic, clinical and job risk factors for these events. The COVID-19 pandemic continues to threat public health, and adverse events frequency and severity among HCWs may get worse. PPE among HCWs is imperative to avoid the widespread diffusion of SARS-CoV-2 but could be harmful due to the long-term utilization. Thus, organizations worldwide should publish guidelines for the appropriate PPE use to prevent these adverse events especially in countries with PPE shortages. Healthcare facilities should take the necessary precautions and change the working conditions during the COVID-19 pandemic (e.g. regular breaks, shorter shifts, adequate supply of PPE, air-conditioning, prophylactic dressing, better material, proper fitting masks, and reduction in wearing time of PPE) to prevent adverse events associated with PPE use and minimize harm to HCWs. Creating a secure and safe work environment for HCWs could lead to a better management of the COVID-19 pandemic and an increase in work performance. Since skin reactions are the more frequent adverse events, policymakers should pay attention to skin hygiene and skin protection including use of skin or sealant protector, protection of injured areas, no use of oily products, wipe of skin to remove sweat, and removal of the masks as frequent as possible. HCWs’ training about appropriate PPE use and knowledge of skin hygiene is of outmost importance. HCWs should recognize symptoms and signs of initial tissue damages adopting then preventive measures to avoid more severe injuries. For example, dry skin and dehydration-induced dermatoses could be avoided with adequate hydration, while moisturizers could help to restore the integrity of skin barrier.

## Data Availability

Data will be available after reasonable request

---

**Web Figure 1.**
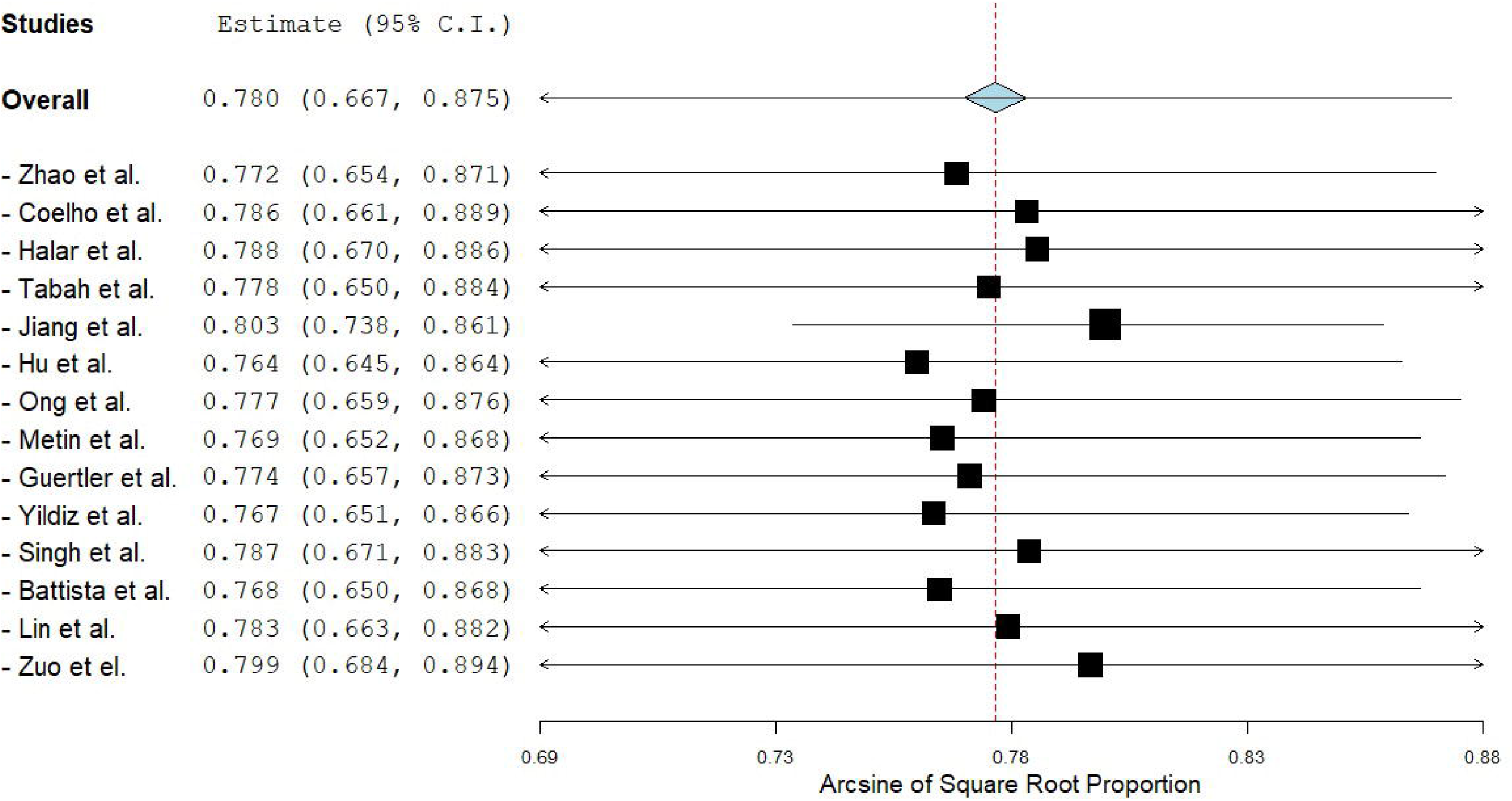
A leave-one-out sensitivity analysis of the prevalence of adverse events among health care workers.

**Web Figure 2.**
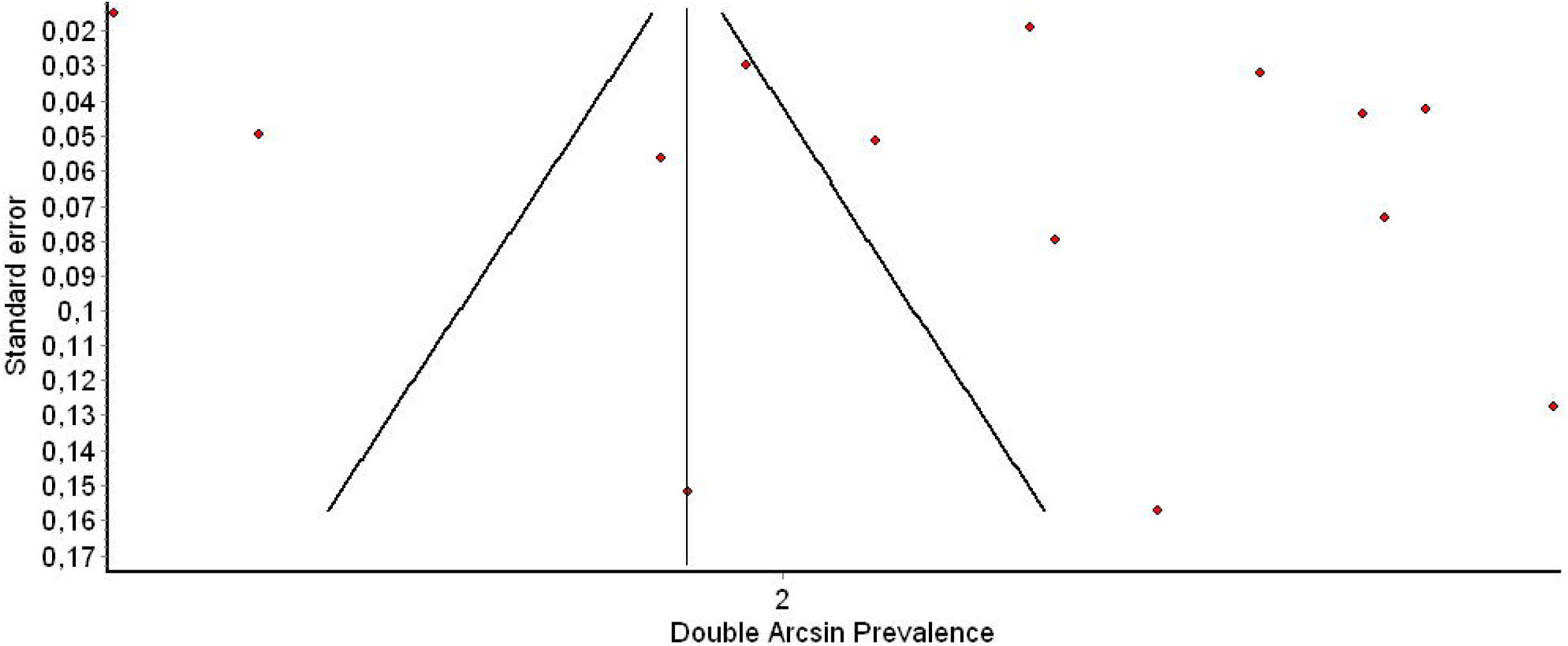
Funnel plot of the prevalence of adverse events among health care workers.

**Web Figure 3.**
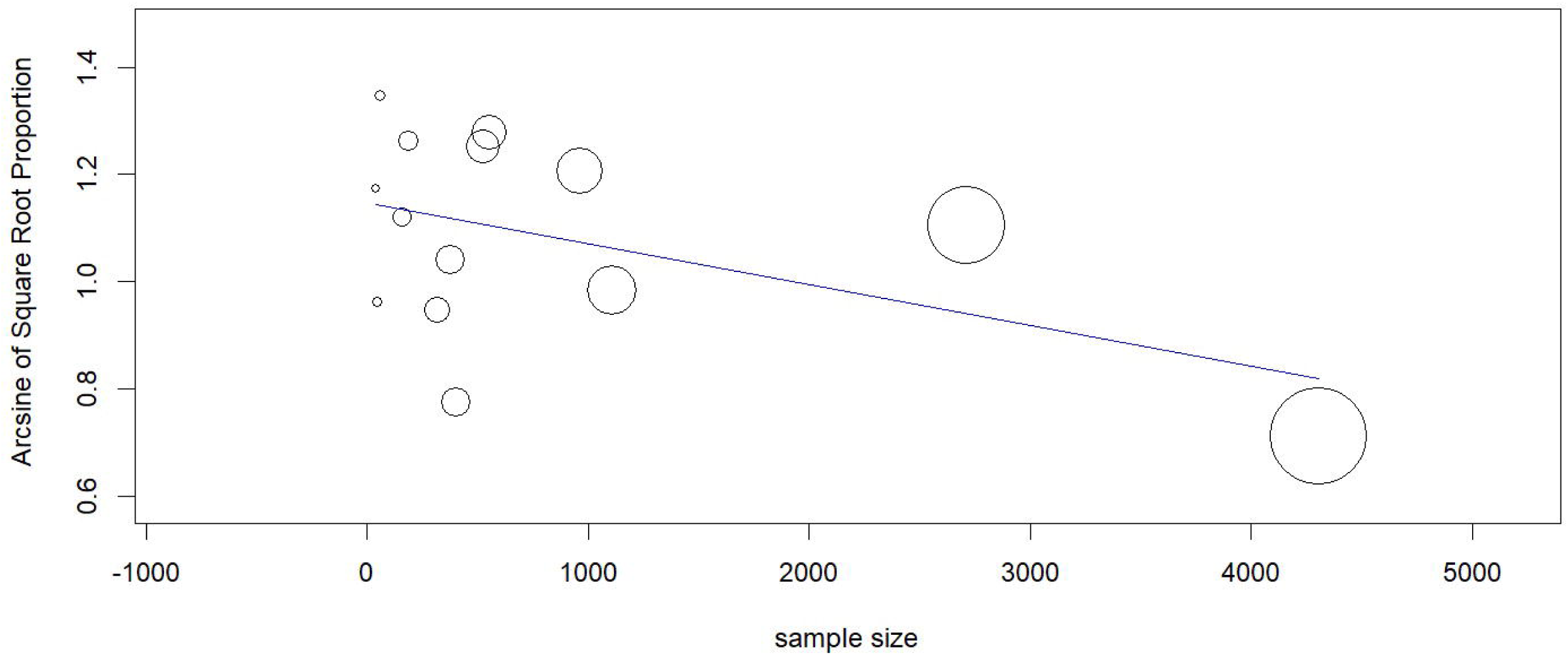
Meta-regression analysis with the prevalence of adverse events among health care workers as the dependent variable and the sample size as the independent variable.

